# Exploring Pain Researcher and Clinician Perceptions of Complementary, Alternative, and Integrative Medicine: A Large-Scale, International Cross-Sectional Survey

**DOI:** 10.1101/2025.02.23.25322738

**Authors:** Jeremy Y. Ng, Dennis Anheyer, Holger Cramer

## Abstract

**Background:** Complementary, alternative, and integrative medicine (CAIM) is gaining popularity among patients experiencing pain, alongside traditional treatments. This study aimed to explore the views of pain clinicians and researchers on CAIM interventions.

**Methods:** An anonymous, online survey was distributed to 46 223 authors who had published pain-related research in MEDLINE-indexed journals. The survey included multiple-choice questions and open-ended sections to gather detailed opinions.

**Results:** A total of 1024 participants responded, most identifying as either pain researchers (43.59%) or both researchers and clinicians (39.88%). Many held senior positions (61.55%). Among the CAIM modalities, mind-body therapies such as meditation, yoga, and biofeedback were viewed as the most promising for pain prevention, treatment, and management, with 68.47% of participants endorsing these approaches. While a majority (43.89%) believed that most CAIM therapies are safe, only 25.55% expressed confidence in their effectiveness. There was broad agreement on the need for more research into CAIM therapies, with 45.88% agreeing and 42.53% strongly agreeing that further investigation is valuable.

Additionally, many respondents supported the inclusion of CAIM training in clinician education, either through formal programs (46.40%) or supplementary courses (52.71%). Mind-body therapies received the most positive feedback, while biofield therapies were met with the most skepticism.

**Conclusion:** These findings highlight the interest in CAIM among pain specialists and emphasize the need for more research and education tailored to this area.

## Introduction

Pain is one of the most common symptoms individuals experience worldwide. Among 146 countries worldwide, 34.1% of individuals were in pain in 2022 [15]. While it is common, the actual experience of pain is subject to great inter-individual variability. Multiple biological and psychosocial variables contribute to these individual differences in pain, including demographic variables, genetic factors, and psychosocial processes. For example, sex, age and ethnic group differences in the prevalence of pain conditions have been widely reported [11,2,6]. Recently the International Association for the Study of Pain (IASP) updated its definition of pain to “an unpleasant sensory and emotional experience associated with, or resembling that associated with, actual or potential tissue damage [13].” Within this definition, there are two primary categories of pain: acute and chronic. Acute pain often presents due to a cause and has a shorter duration, whereas chronic pain is defined as pain that recurs or lasts longer than 3 months [5]. Whether acute or chronic, physicians often turn to a pharmaceutical standard of care when treating sensory or nociceptive pain, with the three most common being acetaminophen, non-steroidal anti-inflammatory drugs (NSAIDs) and opioids. All three of these classes are accompanied with a slew of side effects like nausea, hypertension, abdominal pain and more [32]. As such, there is a need to evaluate cost-effective and lower risk nonpharmacological treatments. This creates a need for patients and clinicians to look towards other forms of medicine to alleviate their pain and discomfort [7].

Complementary, alternative, and integrative medicine (CAIM) has gained increasing popularity over the past decades as a potential treatment option among patients [7]. According to the US National Center for Complementary and Integrative Health (NCCIH), “complementary medicine” is defined as a non-mainstream approach that is used in conjunction with conventional medicine, whereas “alternative medicine” is when the non-mainstream approach replaces the traditional approach [19,22].

Integrative health emphasises a multimodal approach wherein multiple conventional health approaches are utilised alongside complementary approaches [19]. Some common CAIM therapies that are currently used for the management of pain including acupuncture, chiropractic manipulation, massage, and meditation [10]. For example, acupuncture is a practice originating from the East Asian subcontinent that is often practiced to alleviate pain. Despite over 80 systematic reviews being conducted so far to evaluate the impact of acupuncture on pain relief, these reviews have certain degrees of conflict between them, indicating that further research is needed [42]. Chiropractic manipulation is a treatment often sought out by patients experiencing various types of joint related pain. Chiropractors strongly rely on the use of manual therapy, particularly spinal manipulation which is the main form of care they provide [9]. These therapies are often used in conjunction with conventional therapies like pharmaceutical medications and surgery, or as a stand-alone. However, despite the growing popularity of CAIM in pain management, there often is a lack of scientific evidence to support its safety and efficacy [39]. With regards to pain specifically, clinical practice guidelines for a range of CAIMs for the management of pain are uncertain overall, suggesting a need for higher quality recommendations [29,28,27,23]. This presents a clear challenge to clinicians and researchers who are interested in exploring the potential benefits of these therapies.

While some clinicians view CAIM as a valuable addition to conventional healthcare, others remain skeptical of its effectiveness and safety [18]. Thus, there is a clear divide in opinion which has led to varied approaches when integrating CAIM into clinical settings. To understand their perceptions better, we conducted a large-scale, cross-sectional, international study to gather data from pain researchers and clinicians across various geographic regions on their perceptions about CAIM.

## Methods

### Transparency Statement

We sought approval and were granted clearance from the University Hospital Tubingen Research Ethics Board to conduct this study (REB Number: 389/2023BO2). Our protocol was registered on the Open Science Framework (OSF) prior to commencing participant recruitment [31]. All our study materials and data are also available on OSF: https://doi.org/10.17605/OSF.IO/DVECG.

### Study Design

We conducted an anonymous, online, cross-sectional survey consisting of authors who have published articles about or relating to pain in journals indexed by MEDLINE [21].

### Sampling Framework

The sample was determined by searching a series of pain-related MeSH headings and filtering by English-language articles published between May 1, 2016 and May 1, 2023. A complete search strategy can be found in the **Table 1**. The search strategy was run on OVID MEDLINE and the list of PMIDs retrieved was exported. The PMIDs were inputted into an R script to extract the authors’ name, affiliation institutions and emails [33].

**Table 1:** Search Strategy for Author Name and Email Address Retrieval.

|  |  |
| --- | --- |
| **Database: Ovid MEDLINE(R) ALL <1946 to May 23, 2023>**<br>**Search Strategy:** |  |
| **1** | exp Pain, Referred/ or exp Pleasure-Pain Principle/ or exp Pain, Procedural/ or exp Chronic Pain/ or exp Shoulder Pain/ or exp Acute Pain/ or exp Pain Management/ or exp Abdominal Pain/ or exp Pain Measurement/ or exp Nociceptive Pain/ or exp Pain, Intractable/ or exp Visceral Pain/ or exp Pelvic Pain/ or exp Eye Pain/ or exp Pain, Postoperative/ or exp Neck Pain/ or exp Pelvic Girdle Pain/ or exp Labor Pain/ or exp Pain Insensitivity, Congenital/ or exp Low Back Pain/ or exp Pain Threshold/ or exp Pain/ or exp Patellofemoral Pain Syndrome/ or exp Myofascial Pain Syndromes/ or exp Flank Pain/ or exp Chest Pain/ or exp Back Pain/ or exp Facial Pain/ or exp Musculoskeletal Pain/ or exp Pain Clinics/ or exp Complex Regional Pain Syndromes/ or exp Breakthrough Pain/ or exp Pain Perception/ or exp Cancer Pain/ (511661) |
| **2** | limit 1 to english language (434060) |
| **3** | limit 2 to dt=20160501-20230501 (121155) |

### Participant Recruitment

The authors who were identified by utilizing our sampling framework were contacted and asked to complete the survey. Initially, the list of emails went through a screening process where duplicate and/or invalid emails were removed from the dataset. Any names that were extracted incorrectly were corrected; any names that did not have an email associated with them were also removed. The software SurveyMonkey was utilized to send emails to all the authors [36]. This email included an explanation of the study, the study’s goals as well as a link to the survey. Upon clicking the link, participants were initially taken to an informed consent form. The survey began once participants consented to the terms and conditions outlined and indicated their intention to participate. After the initial email was sent on September 04, 2023, subsequent reminders were sent on September 18, 2023, October 04, 2023 and October 18, 2023. After the final reminder was sent, the survey was left open for 4 weeks. Participants could skip any questions; no financial compensation was provided for completing the survey.

### Survey Design

The initial part of the survey consisted of a screening question asking the participant if they were a researcher or clinician within the field of pain, followed by a series of demographic questions to establish certain characteristics of the participant. Patients and the general public were not eligible to partake in the survey. The second part of the survey asked the respondents about their perceptions of CAIM. The survey consisted of 30 multiple choice questions and one open-ended question at the end.

Prior to sending out the survey, it was pilot-tested by two CAIM researchers independently, and their feedback was incorporated prior to distribution. The complete copy of the survey is provided on OSF: https://osf.io/r4cfh.

### Data Management and Analysis

The survey data was analysed and basic descriptive statistics, such as counts and percentages were computed. To analyze the qualitative data, a coding process was performed where similar groups of ideas were assigned a code, which was a concise interpretation of the response provided. Thematic analysis was conducted to narrow down the codes and group similar ones together. The results were reported using the Strengthening the reporting of observational studies in epidemiology (STROBE) checklist [8,40].

## Results

### Search Strategy

After utilizing the search strategy outlined and searching for publications between May 01, 2016 and May 01, 2023, 121 181 articles were extracted along with their accompanying PMIDs were extracted. There were 46 223 unique names and email addresses that were the corresponding authors of the articles. Our survey’s raw, deidentified data is available here: https://osf.io/s3jtu. Crosstabs for key demographic variables can also be found on OSF: https://osf.io/dvecg.

### Demographics

Of the 46 223 emails sent out, 25316 emails were unopened, 14477 were opened and 6430 bounced. A total of 1024 responses were received, thus the response rate for all emails was 2.6%, for received emails (i.e., opened and unopened emails), and 7.1% for opened emails. It is important to acknowledge that not every participant answered all the questions therefore we present the total number of responses for each question in parentheses. Responses were considered incomplete if no answers were provided beyond the initial screening question. The survey took an average of 9 minutes and 47 seconds to complete. Most respondents identified as pain researchers (n=435, 43.59%) or both a pain researcher and clinician (n=398, 39.88%). Some respondents only identified as a pain clinician (n=67, 6.71%) and some responded as neither and were thereby disqualified of the survey (n=98, 9.82%). Respondents were almost equally split among male (n=431, 48.26%) and female (n=443, 49.61%) and were mostly between the ages of 35-44 (n=267, 29.83%) or 45-54 (n=257, 28.72%). Most participants indicated that they were not part of a visible minority group (n=728, 81.43%). With respect to WHO World Regions, participants were primarily located in Europe (n=349, 39.17%) or the Americas (n=318, 35.69%). Respondents also primarily identified as a faculty member/principal investigator (n=448, 50.11%) or a clinician (n=374, 41.83%) in a senior career stage (n=549, 61.55%). When asked to describe their primary research area, most of the respondents stated clinical research (n=571, 73.49%). The characteristics of participants can be found in **Table 2**.

**Table 2.** Characteristics of Survey Participants.

|  |  |
| --- | --- |
| **Self-identification as researcher or clinician (n = 998)** |  |
| Researcher within field of pain | 435 (43.59%) |
| Clinician within field of pain | 67 (6.71%) |
| Both researcher and clinician within field of and pain | 398 (39.88%) |
| **Sex (n = 893)** |  |
| Male | 431 (48.26%) |
| Female | 443(49.61%) |
| Prefer not to say | 13 (1.46%) |
| Prefer not to self-describe | 6 (0.67%) |
| **Age (n = 895)** |  |
| Under 18 | 1 (0.11%) |
| 18-24 | 1 (0.11%) |
| 25-34 | 78 (8.72%) |
| 35-44 | 267 (29.83%) |
| 45-54 | 257 (28.72%) |
| 55-64 | 191 (21.34%) |
| >65 | 91 (10.17%) |
| Prefer not to say | 9 (1.01%) |
| **Visible Minority (n = 894)** |  |
| Yes | 138 (15.44%) |
| No | 728 (81.42%) |
| Prefer not to say | 28 (3.13%) |
| **Location (n = 891)** |  |
| Africa | 22 (2.47%) |
| Americas | 318 (35.69%) |
| Eastern Mediterranean | 40 (4.49%) |
| Europe | 349 (39.17%) |
| South-East Asia | 78 (8.75%) |
| Western Pacific | 75 (8.42%) |
| Prefer not to say | 9 (1.01%) |
| **Current Position (n = 894)** |  |
| Clinician Student | 2 (0.22%) |
| Clinician | 374 (41.83%) |
| Graduate student | 30 (3.36%) |
| Postdoctoral fellow | 76 (8.50%) |
| Faculty member/Principal Investigator | 448 (50.11%) |
| Research support staff (E.g., research manager, research associate, technician) | 38 (4.25%) |
| Scientist in academia | 266 (29.75%) |
| Scientist in industry | 18 (2.01%) |
| Scientist in third sector | 11 (1.23%) |
| Government scientist | 19 (2.13%) |
| Other (please specify) | 38 (4.25%) |
| **Career Stage (n = 892)** |  |
| Graduate or clinician student | 15 (1.68%) |
| Early career researcher (<5 years post education) | 121 (13.57%) |
| Mid-career research (5-10 years post education) | 207 (23.21%) |
| Senior researcher (>10 years post education) | 549 (61.55%) |
| **Primary Research Area (n = 777)** |  |
| Clinical research | 571 (73.49%) |
| Preclinical research (in vivo) | 135 (17.37 %) |
| Preclinical research (in vitro) | 65 (8.37%) |
| Health systems research | 76 (9.78%) |
| Health services research | 160 (20.59%) |
| Methods research | 100 (12.87%) |
| Epidemiological research | 129 (16.60%) |
| Other | 35 (4.50%) |
| **Area of CAIM Research (n = 776)** |  |
| Mind-body therapies | 191 (24.61%) |
| Biologically based practices | 139 (17.91%) |
| Manipulative and body-based practices | 151 (19.46%) |
| Biofield therapy | 18 (2.32%) |
| Whole medical systems | 99 (12.76%) |
| I have never conducted any CAIM research | 324 (41.75%) |
| Other | 62 (7.99%) |
| **Area of CAIM Research (n = 776)** |  |
| Mind-body therapies | 191 (24.61%) |
| Biologically based practices | 139 (17.91%) |
| Manipulative and body based practices | 151 (19.46%) |
| Biofield therapies | 18 (2.32%) |
| Whole medical systems | 99 (12.76%) |
| I have never conducted any CAIM research | 324 (41.75%) |
| Other | 62 (7.99%) |

### CAIM Practices and Experiences

When asked about conducting CAIM research, while some respondents had never conducted CAIM research (n = 324, 41.75%), of those who had, mind-body therapies were the most common research area (n=191, 24.61%). As seen in **Table 3**, participants perceived that mind-body therapies to be the most promising with regards to the prevention, treatment or management of conditions associated with pain (n=569, 68.47%) along with manipulative and body-based practices (n=336, 40.43%). A few participants reported that they did not perceive any CAIM categories to be promising (n=82, 9.87%).

**Table 3:** Perspectives on Most Promising CAIM Categories. CAIM: Complementary, Alternative, and Integrative Medicine.

| **(n = 831)** |  |
| --- | --- |
| Mind Body Therapies | 569 (68.47%) |
| Biologically based practices | 241 (29.00%) |
| Manipulative and Body Based Practices | 336 (40.43%) |
| Biofield Therapies | 63 (7.58%) |
| Whole medical systems | 271 (32.61%) |
| I have never conducted any CAIM research | 82 (9.87%) |
| Other | 52 (6.26%) |

As described in **Table 4**, the CAIM categories for which patients had most often disclosed use were manipulative and body-based therapies (n=320, 76.56%), mind-body therapies (n=297, 71.05%), biologically-based practices (n=277, 66.27%) or whole medical based systems (n=273, 65.31%). When probed about the percentage of their patients who have disclosed or sought to use CAIM interventions, majority of respondents stated either 11-20% (n=86, 20.92%), 0-10% (n=84, 20.44%) or 21-30% (n=71, 17.27%). Over two thirds of clinicians had practiced or recommended mind-body therapies to their patients (n=267, 64.34%) and half of physicians had recommended manipulative and body-based practices to their patients before (n=194, 46.75%).

**Table 4:** Patient Preferences.

|  |  |
| --- | --- |
| **CAIM interventions patients have sought counselling or disclosed using: (n = 418)** |  |
| Mind Body Therapies | 297 (71.05%) |
| Biologically based practices | 277 (66.27%) |
| Manipulative and Body Based Practices | 320 (76.56%) |
| Biofield Therapies | 124 (29.67%) |
| Whole medical systems | 273 (65.31%) |
| I have never conducted any CAIM research | 29 (6.94%) |
| **Percentage of patients who have disclosed using CAIM or seek counseling on CAIM therapies: (n = 411)** |  |
| 0-10% | 84 (71.05%) |
| 11-20% | 86 (66.27%) |
| 21-30% | 71 (76.56%) |
| 31-40% | 42 (29.67%) |
| 41-50% | 32 (65.31%) |
| 51-60% | 26 (6.94%) |
| 61-70% | 15 (3.65%) |
| 71-80% | 19 (4.62%) |
| 81-90% | 10 (2.43%) |
| 91-100% | 26 (6.33%) |
| **Areas that participants have practiced or recommended CAIM to patients: (n = 415)** |  |
| Mind Body Therapies | 267 (64.34%) |
| Biologically based practices | 136 (32.77%) |
| Manipulative and Body Based Practices | 194 (46.75%) |
| Biofield Therapies | 25 (6.02%) |
| Whole medical systems | 125 (30.12%) |
| I have never conducted any CAIM research | 60 (14.46%) |
| Other | 19 (4.58%) |

### CAIM Training and Implementation

As described in **Table 5**, nearly half of the respondents reported not having any formal training in CAIM specialties (n=199, 47.95%). Out of the respondents who received formal training, the top two common fields were manipulative and body-based practices (n=114, 27.47%) and mind-body therapies (n=104, 25.06%). With regards to supplemental training, the responses were varied among categories with most participants having received training in mind body therapy (n=202, 48.91%), manipulative and body-based practices (n=124, 30.02%), biologically based practices (n=120, 29.06%) and whole medical systems (n=105, 25.42%). A similar number of individuals had not received any supplemental training (n=122, 29.54%). Over half of the respondents reported being asked about CAIM occasionally (n=431, 52.05%) while nearly a third of the respondents had been asked about CAIM often (n=269, 32.49%). The remaining respondents reported having never been asked about CAIM (n=128, 15.46%). When asked about where participants would seek out information regarding CAIM, the answer was overwhelmingly through academic literature (n=765, 92.06%) with the next option being through conference presentations or workshops (n=438, 52.71%). Colleagues (n=293, 35.26%) and continuing education courses (n=279, 33.57%) were also among sources that participants would likely seek information from (**Table 6)**.

**Table 5:** Participant Training in CAIM Areas. CAIM: Complementary, Alternative, and Integrative Medicine.

|  |  |
| --- | --- |
| **Formal Training (n = 415)** |  |
| Mind Body Therapies | 104 (25.06%) |
| Biologically based practices | 56 (13.49%) |
| Manipulative and Body Based Practices | 114 (27.47%) |
| Biofield Therapies | 12 (2.89%) |
| Whole medical systems | 53 (12.77%) |
| I have never conducted any CAIM research | 199 (47.95%) |
| Other | 17 (4.10%) |
| **Supplementary Training (n = 413)** |  |
| Mind Body Therapies | 202 (48.91%) |
| Biologically based practices | 120 (29.06%) |
| Manipulative and Body Based Practices | 124 (30.02%) |
| Biofield Therapies | 26 (6.30%) |
| Whole medical systems | 105 (25.42%) |
| I have never conducted any CAIM research | 122 (29.54%) |
| Other | 11 (2.66%) |

**Table 6:** Participant Knowledge Related to CAIM. CAIM: Complementary, Alternative, and Integrative Medicine.

|  |  |
| --- | --- |
| **Members outside of research/clinic asking about CAIM (n = 828)** |  |
| Yes, I have been asked about CAIM often | 269 (32.49%) |
| Yes, I have been asked about CAIM occasionally | 431 (52.05%) |
| No, I have never been asked about CAIM | 128 (15.46%) |
| **Where to seek out CAIM information (n = 831)** |  |
| Academic Literature | 765 (92.06%) |
| Colleagues | 293 (35.26%) |
| Conference presentations or workshops | 438 (52.71%) |
| Continuing education course | 279 (33.57%) |
| Formal clinical training | 160 (19.25%) |
| Government agencies | 142 (17.09%) |
| Health or information pages on the internet | 233 (28.04%) |
| Local or national news | 27 (3.25%) |
| Personal experience | 133 (16.00%) |
| Social media | 41 (4.93%) |
| Working with patients/clients | 171 (20.58%) |
| Other (please specify) | 17 (2.05%) |

### General CAIM Perceptions

In the following questions, respondents were provided with a scale including the following options: “strongly disagree”, “disagree”, “neither agree or disagree”, “agree” or “strongly agree”. When asked about CAIM therapies in general, nearly half of participants agreed with the statement “there is value to conducting research on CAIM therapies” (n=356, 45.88%) with a similar number of participants who strongly agreed (n=330, 42.53%), as shown in **Figure 1**. Similarly, when asked about their level of agreement with the statement “clinicians should receive training on CAIM therapies via formal education” most respondents ranked this statement as “agree” (n=36, 17.74%) or “strongly agree” (n=138, 17.74%). Most of the respondents agreed when asked whether clinicians should receive training via supplementary education as well (n=409, 52.71%). Respondents were split when asked whether they would be comfortable counselling their patients about most CAIM therapies.

**Figure 1:**
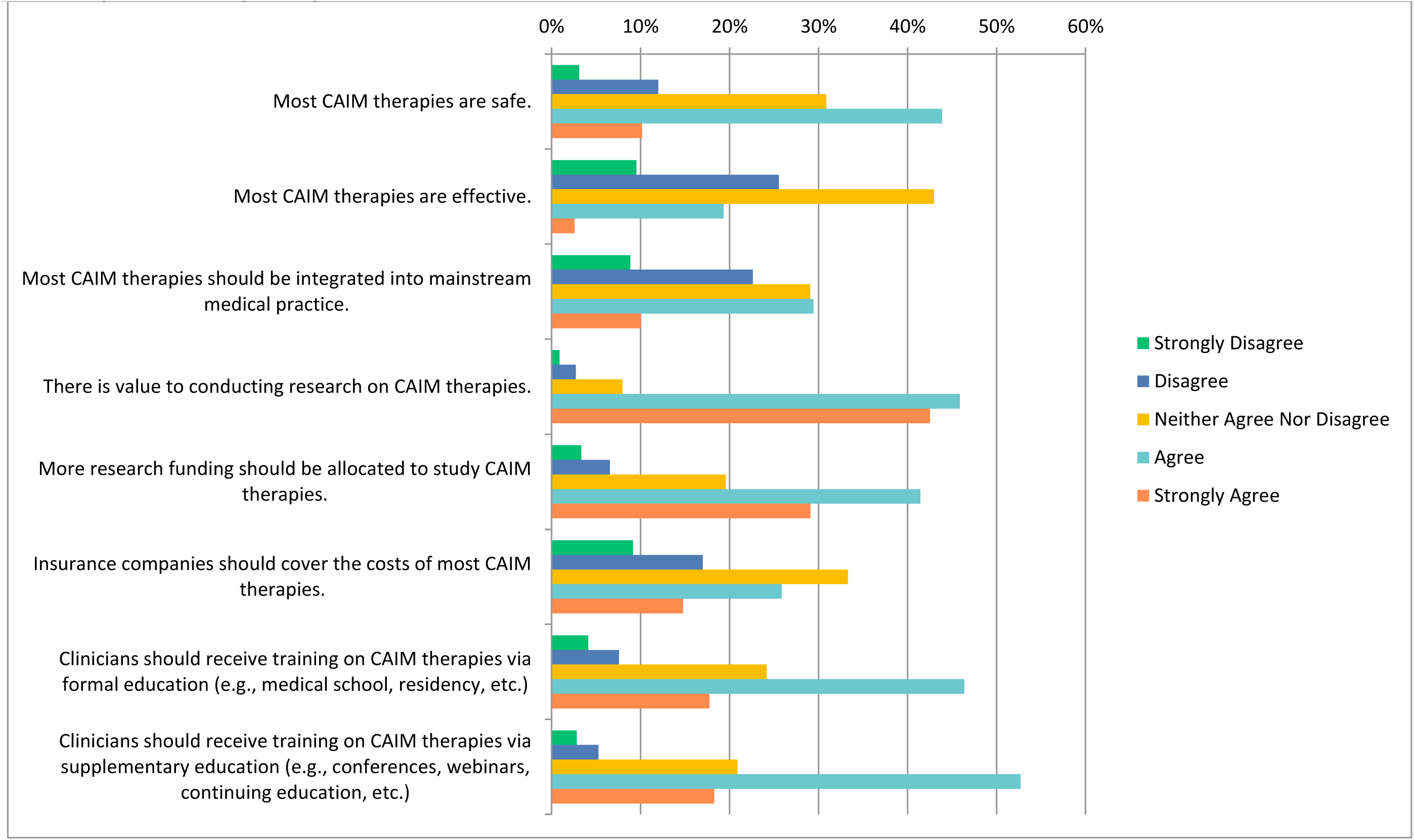
Agreement Regarding CAIM Therapies in General. CAIM: Complementary, Alternative, and Integrative Medicine

Roughly a quarter of patients selected “disagree” (n=100, 25.58%) and another quarter for “neither agree or disagree” (n=101, 25.83%) while a third of patients agreed (n=127, 32.48%). In terms of recommending CAIM to their patients, most of the responses were between “disagree” “neither agree or disagree” or “agree”.

### Specific CAIM Perceptions

The following questions had the same ranking scheme as the previous question set, with references to each specific CAIM category including mind-body therapies, biologically based practices, manipulative and body-based practices, biofield therapies and whole medical systems (**Figures 2-6)**.

**Figure 2:**
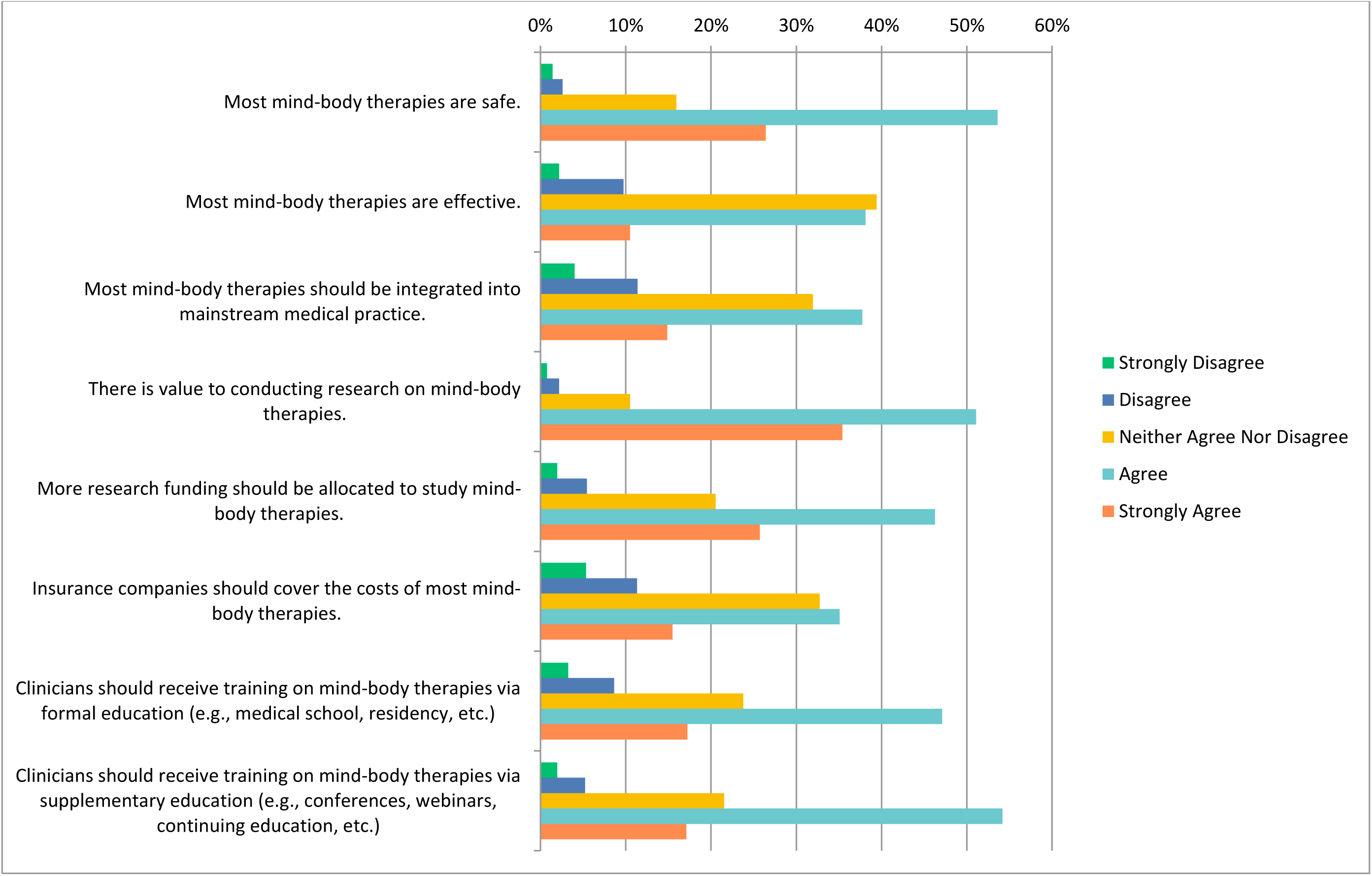
Agreement Regarding Statements about Mind-Body Therapies

**Figure 3:**
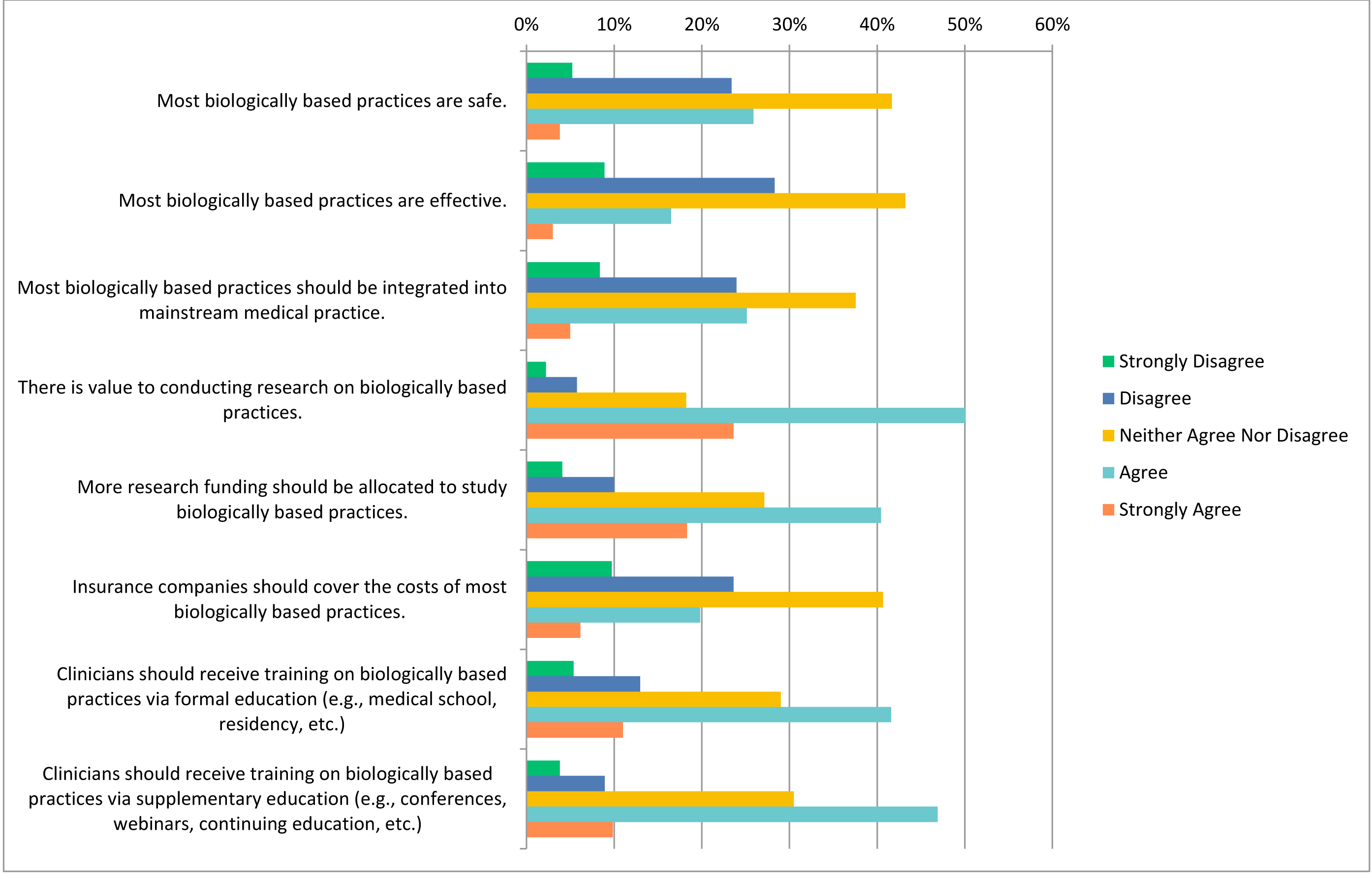
Agreement Regarding Statements about Biologically Based Practices

**Figure 4:**
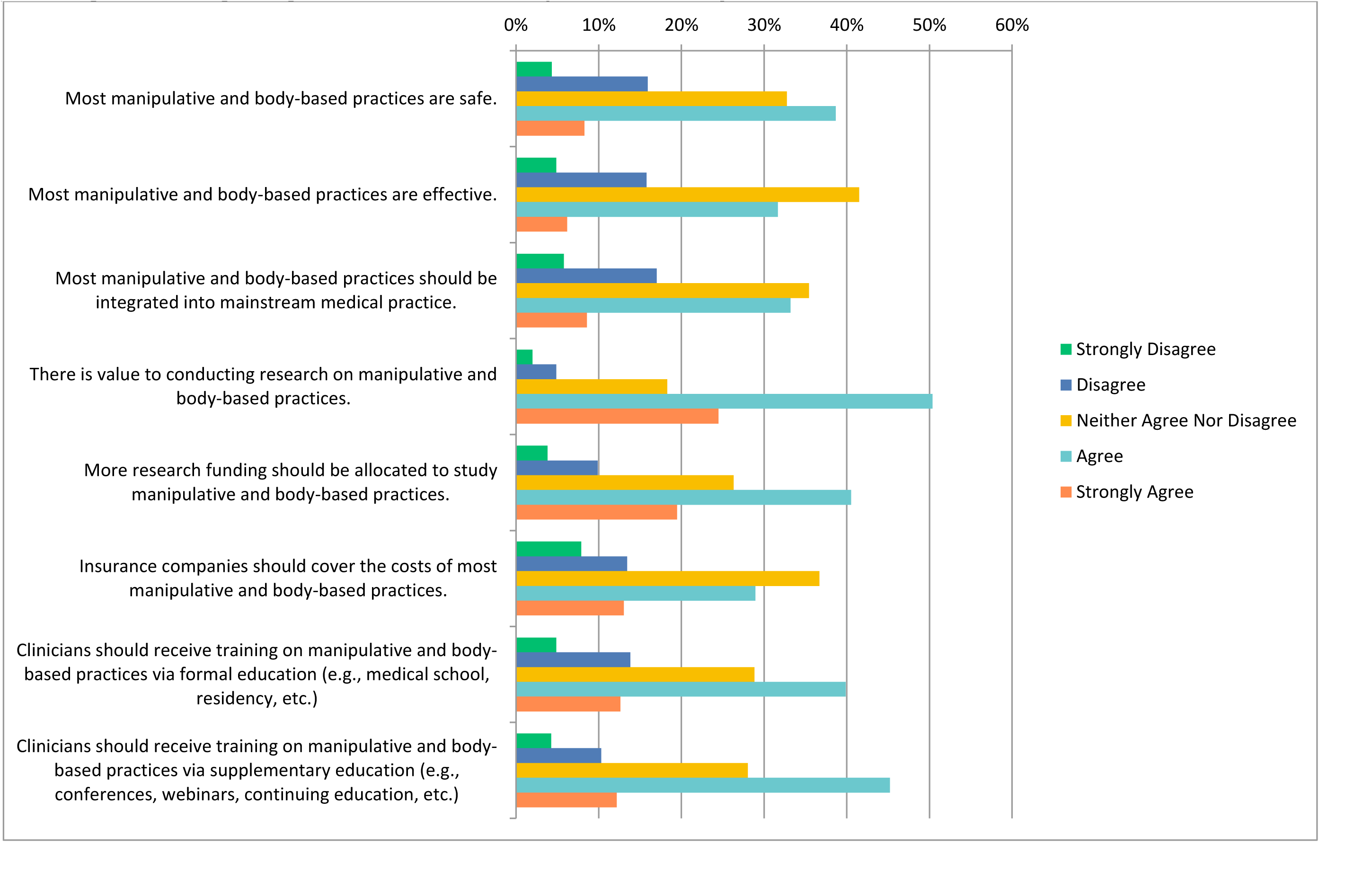
Agreement Regarding Statements about Manipulative and Body-Based Practices

**Figure 5:**
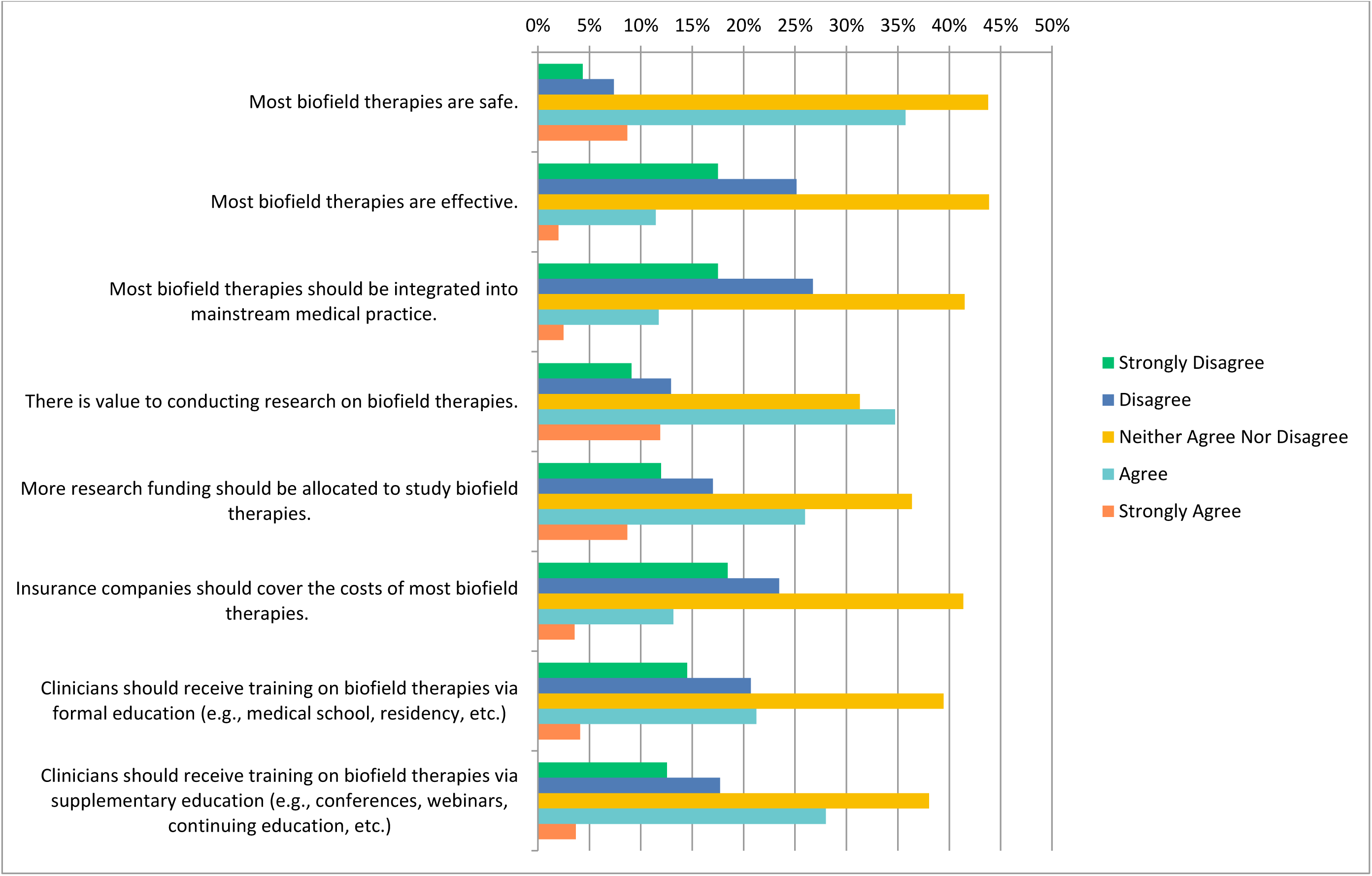
Agreement Regarding Statements about Biofield Practices

**Figure 6:**
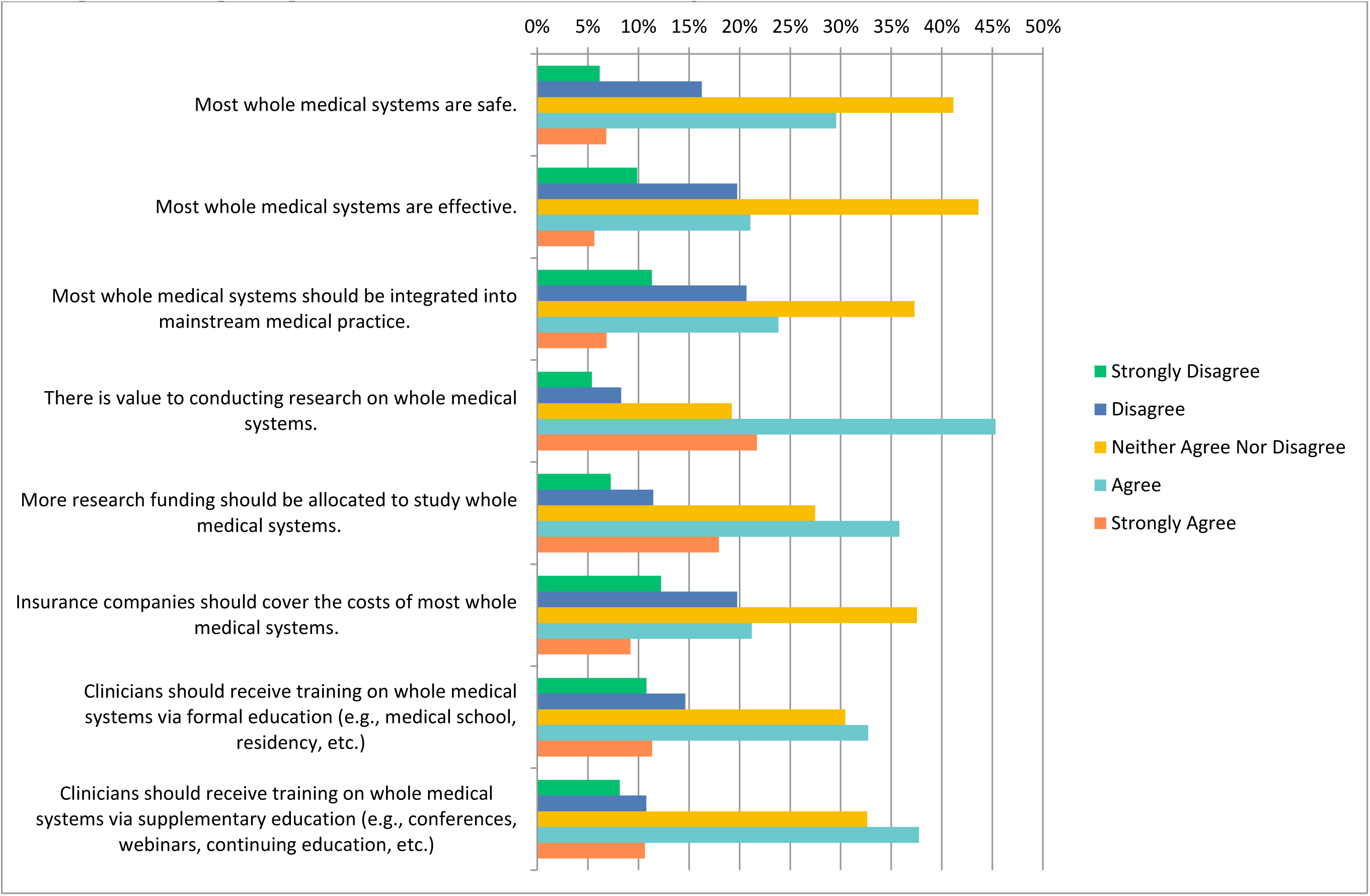
Agreement Regarding Statements about Whole Medical Systems

### Perceptions of Mind-Body Therapy

With regards to mind body therapies (e.g., meditation, biofeedback, hypnosis, yoga, tai chi, imagery, creative outlets), over half of the respondents agreed that most mind-body therapies are safe (n=414, 53.63%) with more than a quarter selected “strongly agree” (n=204, 26.42%). A total of 407/773 individuals either “strongly agreed” or “agreed” with the statement that most mind-body therapies should be integrated into mainstream medical practice. A small portion of individuals either “disagreed” or “strongly disagreed” that there is value to conducting research on mind body therapies (n=23, 2.98%). Almost half of the respondents (n=356, 46%) agreed that more research funding should be allocating to studying mind-body therapies. When asked about receiving mind-body training during clinician training, 361/766 respondents (47.13%) agreed that training should be received via formal education whereas 415 respondents (54.18%) agreed that it should be received via supplementary education (**Figure 2)**.

When asked about counselling patients about most mind-body therapies to patients, 167/391 (42.71%) respondents agreed with 51 disagreeing (13.04%). In terms of recommending mind-body therapies to patients, 40.36% of respondents selected “agree”, 21.85% selected “neither agree nor disagree” and 16.45% selected “disagree”.

### Perceptions of Biologically Based Practices

Around 41.70% neither agreed nor disagreed on the safety of these practices, while 43.25% agreed that most biologically based practices are effective. Regarding integration into mainstream medical practice, the opinions were more varied as 37.57% neither agreed nor disagreed and 25.13% agreed. The majority (50.13%) believed that there is value in conducting research on biologically based practices, and a significant proportion (40.45%) agreed that more research funding should be allocated to studying them. However, opinions on insurance coverage were mixed, as 40.68% neither agreed nor disagreed, while 23.62% disagreed. With regards to clinician training, there was support for both formal and supplementary education as 41.60% agreed that clinicians should receive training through formal education and 46.91% endorsed supplementary education such as conferences and webinars **(Figure 3)**.

When asked about counseling patients about biologically-based therapies, 29.46% of respondents (n=114) agreed, while less than a quarter of respondents disagreed (n=96, 24.81%). Around a third of respondents disagreed with recommending biologically based practices (n=125, 32.30%) whereas a fifth agreed with recommending such practices (n=79, 20.41%). Additionally, 11.63% (n=45) strongly disagreed with counseling, and 13.95% (n =54) strongly disagreed with recommending biologically based practices.

### Perceptions of Manipulative and Body-Based Practices

294/760 participants (38.68%) strongly agreed that most manipulative and body-based practices are safe, while a proportion of 31.67% (n=241) agreed that these practices are effective. Concerning integration into mainstream medical practice, respondents were divided, with 35.44% agreeing and 33.20% disagreeing.

Additionally, half of the respondents (n=383, 50.39%) believed that there was value in conducting research about such practices. A notable number (40.53%) thought that more research funding should be allocated to studying these practices. Opinions on insurance coverage varied, with 36.71% neither agreeing or disagreeing that insurance companies should cover the costs, while 28.95% agreed and 13.42% disagreed. Regarding clinician training, more than a third of respondents (n=303, 39.87%) agreed that clinicians should receive training through medical school or residency, with another 12.67% (n=96) strongly agreeing. With regards to supplementary education, like conferences and webinars, 45.24% of participants agreed and 12.17% strongly agreed **(Figure 4)**.

When asked about whether respondents would counsel their patients regarding manipulative and body-based practices, 56 strongly agreed (14.55%), 136 agreed (35.32%), 94 neither agreed or disagreed (24.42%), 74 disagreed (19.22%) and 25 strongly disagreed (6.49%). The results were more divided when asked about recommending biologically based practices to their patients. A quarter of individuals disagreed (n=98, 25.45%), another quarter of individuals neither agreed nor disagreed (n=95, 24.68%) and around 28.05% of individuals agreed (n=108).

### Perceptions of Biofield Therapies

More than a third of participants (n=271, 35.75%) believed that most biofield therapies are safe, while a large proportion of participants (n=332, 43.80%) neither agreed nor disagreed. Opinions with regards to the effectiveness of biofield therapies were more negatively perceived with a quarter of participants disagreeing (n=191, 25.16%) and 17.52% of individuals strongly disagreeing (n=133). When asked about the integration of biofield therapies into mainstream practices, 133 of 759 respondents strongly disagreed (17.52%), over a quarter of respondents disagreed (n=203, 26.75%) and less than half of individuals neither agreed nor disagreed (n=315, 41.50%). Regarding the value of research, a portion of 34.74 (n=263) agreed that there is value in conducting research on biofield therapies, while 31.31% (n=237) neither agreed nor disagreed. Additionally, a quarter of participants (n =197, 25.96%) agreed that more research funding should be allocated to study biofield therapies. In terms of insurance coverage, opinions varied with 23.45% of individuals disagreeing that that insurance companies should cover the costs of most biofield therapies (n=178) and 41.37% neither agreeing nor disagreeing (n=314). Regarding clinician training, there were similar results for both formal and supplementary education **(Figure 5)**.

The majority of participants either strongly disagreed (n=76, 19.84%) or disagreed (n=119, 31.07%) with the statement that they would be comfortable counselling patients about most biofield therapies. Similarly, for recommending these therapies, over a quarter (n=102, 26.63%) strongly disagreed or disagreed (n=125, 32.64%).

### Perceptions of Whole Medical Systems

Notably, 41.13% of participants expressed neutrality on the safety of most whole medical systems, while a similar percentage (43.61%) neither agreed nor disagreed with their effectiveness. Regarding integration into mainstream medical practice, a significant portion (37.29%) neither agreed nor disagreed. Participants acknowledged the value of research on whole medical systems, with 45.32% agreeing, and 35.80% of respondents agreed with increased research funding. On the matter of insurance coverage, 37.55% neither agreed nor disagreed and were neutral. In terms of clinician training, 32.72% agreed that formal education should include whole medical systems, while 37.76% agreed that it should be within supplementary education (**Figure 6)**.

For counseling, 28.24% neither agreed nor disagreed, while 24.35% agreed. In contrast, 15.28% strongly disagree with counseling. Regarding recommendations to patients, 28.31% neither agreed nor disagreed, while similar percentages of participants agreed and disagreed. A smaller percentage, 6.49%, strongly agreed with recommending whole medical systems to patients.

### Perceived Benefits and Challenges

When asked about the benefits associated with CAIM, 556 out of 748 respondents (74.33%) stated “expanded treatment options for patients”. The second and third most voted options were “holistic approach to wellness” (n=507, 67.78%) and “empowerment of patients to take control of their own health” (n=477, 63.77%) respectively. Of the 46 respondents (6.15%), who provided their own response under “other”, the common themes that emerged were 1) there are no benefits of CAIM, 2) CAIM is effective in reducing pain and 3) reduced medication use **(Table 7)**.

**Table 7:** Benefits and Drawbacks Associated with CAIM. CAIM: Complementary, Alternative, and Integrative Medicine.

|  |  |
| --- | --- |
| **Benefits (n = 748)** |  |
| Expanded treatment options for patients | 556 (32.49%) |
| Holistic approach to health and wellness | 507 (52.05%) |
| Empowerment of patients to take control of their own health | 477 (15.46%) |
| Fewer side effects than conventional medicine | 326 (43.58%) |
| Increased patient satisfaction and well-being | 391 (52.27%) |
| Focus on prevention and lifestyle changes | 459 (61.36%) |
| Cultural and spiritual relevance for certain populations | 379 (50.67%) |
| Potential cost savings for patients and healthcare systems | 278 (37.17%) |
| Potential to address chronic health conditions that conventional medicine has been unable to treat effectively | 403 (53.88%) |
| Integration with conventional medicine to provide CAIM options | 361 (48.26%) |
| Other (please specify) | 46 (6.15%) |
| **Drawbacks (n = 755)** |  |
| Lack of scientific evidence for safety and efficacy | 661 (87.55%) |
| Lack of standardization in product quality and dosing | 627 (83.05%) |
| Limited regulation and oversight | 522 (69.14%) |
| Limited integration with mainstream healthcare systems | 430 (56.95%) |
| Stigmatization and skepticism from healthcare providers and the public | 346 (45.83%) |
| Limited availability in certain geographic areas or for certain populations | 298 (39.47%) |
| High cost and lack of insurance coverage | 370 (49.01%) |
| Potential interactions with prescription medicine | 325 (43.05%) |
| Limited patient education and understanding | 352 (46.62%) |
| Difficulty in distinguishing legitimate practices from scams or fraudulent claims | 532 (70.46%) |
| Other (please specify) | 43 (5.70%) |

Additionally, participants were asked about challenges that they perceive are associated with CAIM. Out of 755 participants, 661 (87.5%), believed that the lack of scientific evidence for safety and efficacy was the biggest challenge. Following closely behind as the highest voted challenge was lack of standardization in product quality and dosing (n=627, 83.05%) and limited regulation and oversight (n=522, 69.14%). When asked to further share their opinions via an “other option”, 43 participants (5.70%) provided further thoughts with varying themes. Some of the common themes that emerged were misrepresentation of effectiveness, lack of standardization and risk of false hope (**Table 7)**.

### Thematic Analysis

When respondents were asked if they had anything else to share about their perceptions of CAIM, 797 individuals skipped the question however 236 individuals answered the question with a range of responses. In total, 23 codes were identified from 155 open-ended responses. These codes were then summarized and grouped further to create 7 distinct themes – which are specific patterns that were found in this dataset. Firstly, “concerns with CAIM categorization” included individuals who disagreed with the grouping of CAIM modalities in the survey and believed that some categories were too broad or too narrow. Secondly, “CAIM is effective if beneficial to patient and integrated into practice” included individuals who believed that efficacy was of foremost concern, and that patient results matter most. Another category was “CAIM should be studied using evidence-based methods”, which included responses that advocated for further research and for rigorous scientific evidence. The next category was “polarized opinions about CAIM” which included individuals who were either for or entirely against CAIM modalities being integrated into treatment. Finally, “physicians require CAIM education” encompassed responses that described the need for further education in various levels of institutions regarding CAIM. Coding and thematic analysis data are available at: https://osf.io/d45z6.

## Discussion

The purpose of this study was to explore the perceptions of pain researchers and clinicians about CAIM. Participants found mind-body therapy to be the most promising CAIM category and the area that the clinician/researchers had practiced most frequently or recommended to patients. On the contrary, participants were more sceptical about biofield therapies and biologically based practices. Participants also agreed that conducting further research on mind-body therapies and manipulative and body based practices is the most valuable, and that clinicians should receive training on CAIM therapies via formal or supplementary education.

### Comparative Literature

The conclusions from our survey are consistent with other literature regarding clinicians’ and researchers’ perceptions of CAIM from various medical specialties. In one similar survey, it was found that oncology researchers and clinicians perceived mind-body therapies to be the most effective and safe CAIM modality. It was also found that oncology researchers and clinicians believed that more CAIM training should be provided via formal or supplementary education [25]. Similarly, a study focused on neurology researchers and clinicians found that 82% of respondents believed that there was value in conducting CAIM research and 58.9% believed that more funding should be allocated towards this research [26]. The most favoured CAIM modality was also mind-body therapies in this study, with the two least favourable being biofield therapies and whole medical systems. This study found that a lack of scientific efficacy and safety was reported to be the greatest challenge to CAIM. Moreover, a study that investigated psychiatry clinicians’ and researchers’ perceptions of CAIM also found mind-body therapies to be the most promising CAIM therapy for the management of psychiatric diseases [24].

In our study, mind-body therapies (e.g., yoga, meditation, biofeedback, tai chi) were considered the most promising with regards to prevention, treatment, and management of pain, and biofield therapies received the most negative responses in the study. This is consistent with other studies conducted within the field of pain. One study that explored physicians, nurses, physical therapists, and midwives’ perceptions of complementary medicine for chronic pain found similar results. Yoga and tai chi were among the most positively perceived modalities while reiki and biofeedback were modalities that participants possessed the least familiarity [1].

Similarly, in a survey conducted in the UK, physiotherapists perceived acupuncture, massage, osteopathy, and yoga to be effective in the treatment of lower back pain yet were sceptical about other modalities like reiki, shiatsu and herbal medicine [12]. Another study investigated health-care practitioners’ perceptions of yoga and found a high likelihood of the physicians recommending yoga to their patients for musculoskeletal pain and mental health concerns [35]. Another found that physicians were most likely to recommend acupuncture to patients who suffered from migraine related pain [30]. Further, a study of pain fellowship directors at the American College of Graduate Medical Education found that many directors reported acupuncture as a modality they frequently discuss with patients in a chronic pain treatment plan [16]. With regards to researchers’ perceptions, a bibliometric analysis of meditation trends revealed that researchers continue to trust the clinical benefits of mindfulness meditation in pain management [34]. On the contrary, biofeedback therapies like reiki might have been perceived negatively as they are most prevalent in Asia [17], and most of our respondents were based in Europe or the Americas.

The findings from our survey are consistent with previous literature surrounding clinicians and their perceptions of CAIM. In our study, participants agreed there is value in conducting research on CAIM, and that further funding should be allocated to it. As our study primarily sampled researchers or researchers and clinicians, this could potentially explain why they would believe in conducting further research. Further, participants agreed that there is value in providing formal or supplementary CAIM training to clinicians, which is consistent with literature concerning psychiatrists, oncologists and neurologists [25,26,24]. The participants sampled in our study were mostly senior researchers and clinicians, who might have completed their training when CAIM therapies were not as prevalent in the healthcare landscape, and thus would support its integration into medical training curricula. The thematic analysis we conducted also revealed that participants found the lack of research to be the biggest barrier to their perceptions on CAIM, and that only few modalities are backed with evidence, suggesting a need for future research.

The two major benefits that were reported, were expanded treatment options and a holistic approach to health and wellness. Expanded treatment options was likely ranked highest as pain is often a multidimensional condition with complex physical and psychological comorbidities [14]. CAIM being a holistic approach was also viewed positively, and is concurrent with previous studies, where physicians believed strongly in the holistic nature of complementary and alternative medicine [4,38]. With regards to challenges, one of the biggest challenges that was reported was a perceived lack lack of scientific efficacy for the modalities, which is consistent with a paper that found that one of primary care providers’ largest barrier was scepticism of efficacy [3].

### Strengths and Limitations

The study generalized researchers’ and clinicians’ perceptions of CAIM due to the large and international random sample of individuals possessing varying opinions on the topic. The email addresses and names of participants were found using MeSH headings Medical Subject as it is an effective way of categorizing articles [20]. To maximize the response rate, we sent out multiple reminders via email throughout the survey duration. We only sent emails to individuals who have published over the last seven years, thereby limiting the potential for inactive addresses and maximize response.

A key limitation of the study is that it is likely that we only sampled participants who understand English, as we only sampled participants who had written journal articles in English and the survey itself was in English. Our response rate is likely underestimated due to certain email addresses being inactive or invalid; additionally, some individuals were potentially on vacation, out of office, retired, or have passed away and were unable to respond. There were also likely many non-responding participants who received the email but did not complete the survey, which is often an inherent limitation of collecting data utilizing a survey. Nonresponse bias could have occurred when the characteristics of the non-responders vary from the responders [37]. There could have been a potential for recall bias as participants might have struggled with recounting certain information. The study was also susceptible to sampling bias as we are selecting subjects from a large population [41].

## Conclusions

Overall, participants had varying opinions when it came to the adoption of specific CAIMs into their practices, with mind-body therapy and manipulative and body-based practices being the most well perceived, however there was a clear indication for a need for further research. To the best of our knowledge, this is the first study to document how pain clinicians and researchers view CAIM and its benefits/drawbacks. The data and analysis presented within this paper can potentially guide future research surrounding CAIM, and its use in the field of pain research and pain treatment.

## List of Abbreviations

CAIM: complementary, alternative, and integrative medicine
IASP: International Association for the Study of Pain
MeSH: Medical Subject Headings
STROBE: Strengthening the reporting of observational studies in epidemiology

## Data Availability

All study materials and data are included in this manuscript or posted on Open Science Framework.

https://doi.org/10.17605/OSF.IO/DVECG

## Acknowledgement Section

Ethics Approval and Consent to Participate

We sought and were granted ethics approval by the University Hospital Tubingen Research Ethics Board to conduct this study (REB Number: 389/2023BO2).

## Consent for Publication

All authors consent to this manuscript’s publication.

## Availability of Data and Materials

All study materials and data are included in this manuscript or posted on Open Science Framework: https://doi.org/10.17605/OSF.IO/DVECG

## Authors’ Contributions

JYN: designed and conceptualized the study, collected and analysed data, drafted the manuscript, and gave final approval of the version to be published.

DA: assisted with the analysis of data, made critical revisions to the manuscript, and gave final approval of the version to be published.

HC: assisted with the analysis of data, made critical revisions to the manuscript, and gave final approval of the version to be published.

